# Resilience Assessment of healthcare facilities within urban context: Learning from a non-profit hospital in Tehran, Iran

**DOI:** 10.1101/2021.09.13.21263435

**Authors:** Mehrdad Rahmani, Aynaz Lotfata, Sarasadat Khoshnevis, Komar Javanmardi

## Abstract

Tehran’s healthcare system is under increasing strain due to population expansion and a lack of disaster preparedness measures. The purpose of this paper is to analyze hospital resilience in an urban setting to identify areas for improvement to keep the studied hospital operational during a crisis. In this study, the Urban Resilience Index (URI) in Amir-Alam hospital was assessed using a customized version of the City Resilience Profiling Tool (CRPT) established by the United Nations Office for Disaster Risk Reduction (UNISDR). The 36 indicators were analyzed in 5 components. The result has revealed that the hospital’s urban resilience score was calculated to be 51.75 out of 100, indicating medium resilience, while, regarding the critical indicators, the score was 20.25 out of 60, which is not acceptable. The physical, organizational, and spatial attributes of the Amir-Alam hospital are among the least resilient, but the functional and dynamic characteristics are reasonably decent.

## Introduction

Urban systems are complex networks of interdependent subsystems [**1**]. Cities’ adaptation to urban outbreaks relies on the mutual exchange of services of various intertwined and interdependent systems and subsystems, for instance, healthcare, energy, transport, manufacturing, and financial departments [**2, 3**]. Urban resilience and its assessment have become an area of interest to provide a lens to understand the complexities of cities as socio-ecological systems. In contrast, extreme events such as pandemics, earthquakes, and floods have revealed long and expensive recovery processes of affected cities [**4**]. The pre-disaster resilience assessment and mitigation actions and smart and strategic urban planning in disaster-prone regions reduce loss and damages [**5**].

Natural and human-made disasters impose a range of damages and losses to the affected societies. Injuries are associated with the infrastructure and facilities, such as hospitals and emergency services. Healthcare services are on the front line of disaster response. They must remain operational regardless of the community context and level of preparedness to provide needed services to an affected population. Unexpected disasters affect service providers such as hospitals during catastrophic situations; the community role extends beyond a structural entity that offers healthcare services. Markedly, hospital safety from disasters is a challenge in all countries **[6, 7]**, while there has been less attention to the comprehensive pre-hazard assessment of hospital resilience **[8, 9]**. In recent years, disasters resulted in hospital damages and interruptions in medical services **[10, 11]** as a devastating earthquake in Ezgele, Kermanshah, on the west side of Iran in November 2017 **[12]**. That is why, in 2015, at the third world conference held in Sendai-Japan, the resilience of health infrastructures and disaster risk reduction measures were highlighted **[13]**.

Hospital resilience defines as a hospital’s capability to resist, absorb, and respond to the shock of disasters while still retaining its most essential functionality (e.g., prehospital care, emergency medical treatment, critical care, decontamination, and isolation), then recover to its original state or a new adaptive state. More specifically, hospital resilience is the capability to absorb the impact of disasters without loss of functions (termed resistance); maintain its most essential functions (called absorption and responsiveness), and bounce back to the pre-event state (termed recovery) or to a new form of operation (termed adaptation) [**14**]. Resilience asset management cannot be dealt with in the isolation of individual sectors but should be seen holistically across multiple sectors of urban resilience. Hence, we need to identify how we assess our assets and manage them in the case of city preparedness through urban resilience enhancement. The asset management decisions should be made across multiple sectors and in line with the critical infrastructures’ resilience plan [**15**].

Hospital resilience is an emerging concept that addresses the crucial issues around reducing vulnerabilities and enhancing city stability. There is a worldwide significant amount of research to quantify hospital resilience like the Hospital Safety Index (HSI) to assess the safety of hospital buildings, critical systems, and equipment, the availability of supplies, and the emergency and disaster management capacities [**16, 17**]. The other range of studies examined the hospital resilience on the operational function of the hospitals in terms of patients’ satisfaction and patient waiting time [**18**], economic resilience of hospitals [**19**], architectural resilience in facing environmental crises like earthquakes and floods (e.g., [**6, 20, 21, 22, 23**]). While these studies contribute to assessing the building safety level and hospital capacity in emergency conditions, they do not address the hospital resilience in terms of the urban context’s resilience within which it is located. For example, Zhong proposed multiple variables for assessing hospitals’ resilience in China, including hospital safety, emergency services, surge capacity, command, disaster plan, logistics, staff ability, disaster training, communication and cooperation systems, recovery, and adaptation [**24**].

Multi-scaled understanding of hospital resilience embraces operational and non-operational aspects and structural and non-structural criteria to assess resilience within the hospital building and its surrounding urban context. Additionally, resilience management emphasizes several interdependent urban components that operate as one system at multiple (local, metropolitan, regional, national, global) scales [**25**]. Therefore, it is prominent to encourage researchers, practitioners, and decision-makers to develop a multi-scale approach to hospital resilience.

The overall aim of this study is the multi-dimensional and multi-scalar assessment of hospital resiliency in an urban context and finding the area of improvement in the hospital resilience for maintaining its function in the disaster period. So, the hospital building resilience has been overlooked in terms of the significance of urban context resilience that surrounds it.

### Multi-Dimensional Resilience Assessment

The way cities and communities dealing with natural and human-made outbreaks shows the extent of urban resilience. Urban areas are vulnerable to these outbreaks due to their higher population, densities, and mobility. That is why urban resilience is associated with Resilient Urbanism that addresses the resilient urban form, urban development, and urban management [**26**]. Accordingly, it should be highlighted that urban resilience is an integrative concept that encourages the resilient management of urban assets. In that way, it increases the public safety and well-being of people during urban outbreaks.

Likewise, vulnerability to disasters varies geographically [**27, 28**], and resiliency does not merely address a physical aspect. Another aspect’s role is in that such as institutional, social, environmental, infrastructure, and economic aspects. Jabareen [**29**] stated that a resilient city defines by the overall abilities of its governance, physical, economic, and social systems. Entities exposed to hazards to learn, be ready in advance, plan for uncertainties, resist, absorb, accommodate to and recover from the effects of the threat in a timely and efficient manner through the preservation and restoration of its essential basic structures and functions [**29**]. Therefore, multi-dimensional spatial management is prominent for a better overview of mobility, spatial networks, socio-spatial dynamics, functionalities, and spatial arrangements. For instance, the study of earthquake dynamics offered the need for spatial analysis to evaluate health care system safety and accessibility [**10**]. Urban asset management needs the action plan at multiple scales as urban resilience is a multi-faceted aspect. Ideally, all different dimensions of an urban system should be addressed in a resilience assessment framework.

The issue of resilience is part of our life as we deal with natural and human-made hazards. In the urban context, the resilience enhancement usually occurs through a mitigation/contingency plan that offers suggestions for keeping safe access, adequate public services, and responding to any vulnerabilities in the city. It requires the integrated function of multiple scales in spatial management and multi-dimensional assessing, planning, and development of spatial profiles to enhance the adaptive capacity of urban assets. In other words, to strengthen the resilience of urban investment, we must address the structural and functional attributes of the hospital and the specific factors of surrounding urban systems, multiple sectors, governance, and management.

Management of urban systems could study from the perspective of Urban Resilience Policy (e.g., [**30**]). There is an ongoing need for urban resilience evidence-gathering processes to be multi-scalar and multi-dimensional, ideally advancing the resilience assessment framework. Those mobilize a wide range of related stakeholders into a collective and collaborative effort where technical elements of resiliency are fused with social and organizational requirements. In addition, it should be highlighted the UN-Habitat six characteristics of persistent, adaptable, inclusive, integrated, reflexive, and transformative urban resilience characteristics in urban hazard studies [**31**].

## Data and Method

### Case Study

The Amir-Alam hospital is located in the old city center (Mantaghe 12) of Tehran. It is delaminated by Memar-Makhsuos street (North), Saadi Street (East), Taghavi street (South), and Lalehzar street (West). This hospital complex consists of 3 different sets of buildings: the old site of the hospital and two new buildings on the north and south sides of the old hospital. It is a 10500 m2 hospital with 196 beds and almost 800 staff **(Figure 1)**.

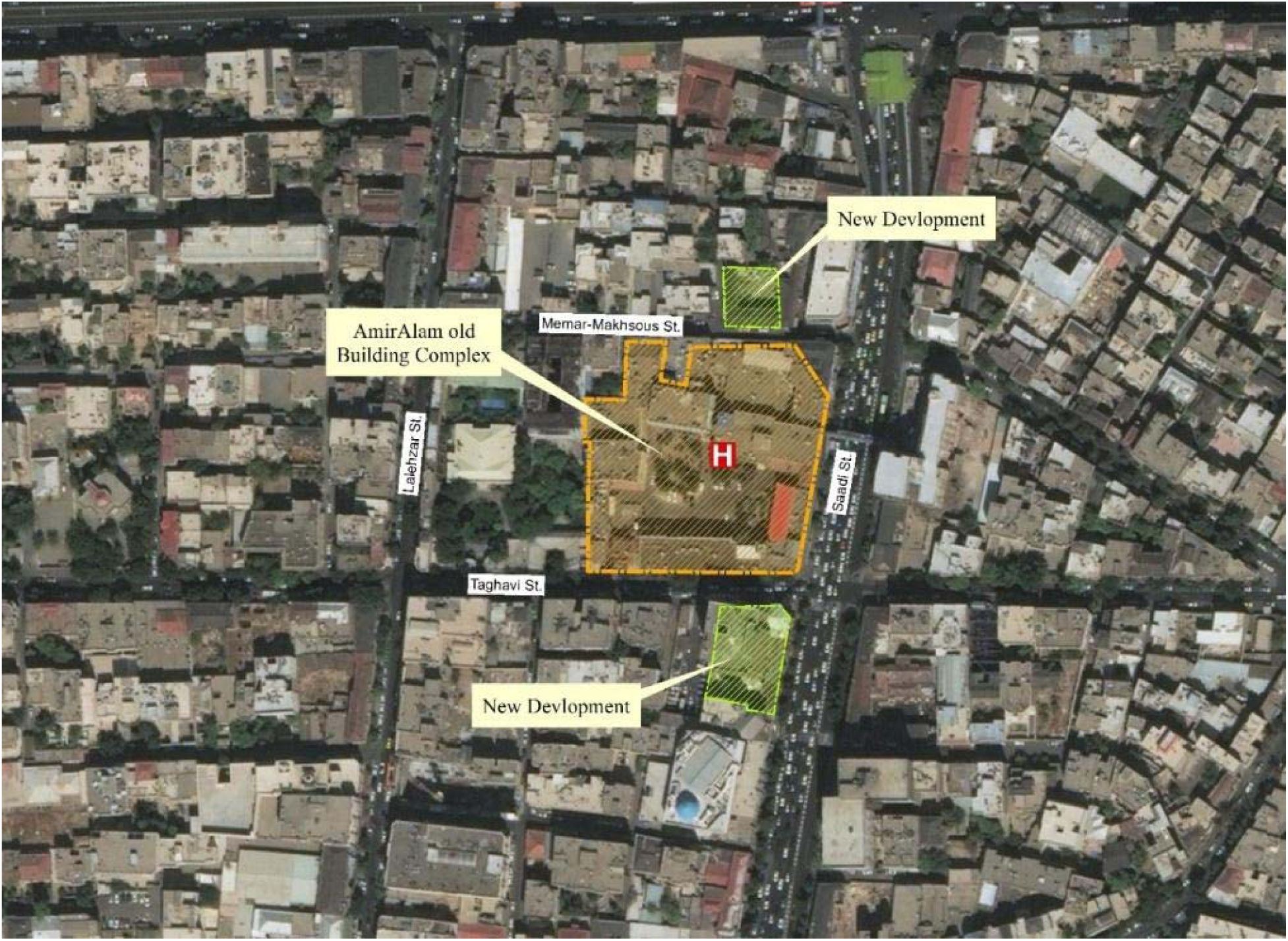

Four spatial zones surrounding Amir-Alam hospital defined to conduct the multi-scalar assessment (Figure 2):

- Structural zone including the site of the hospital and the closest spaces;
- Adjacent zone including walkable service area of the hospital calculated by network analysis in ArcMap 10.2;
- Functional zone including the total service area of the hospital that and
- District zone, including the district 12 of Tehran which is an administrative unit of the city.

**Figure 2.**
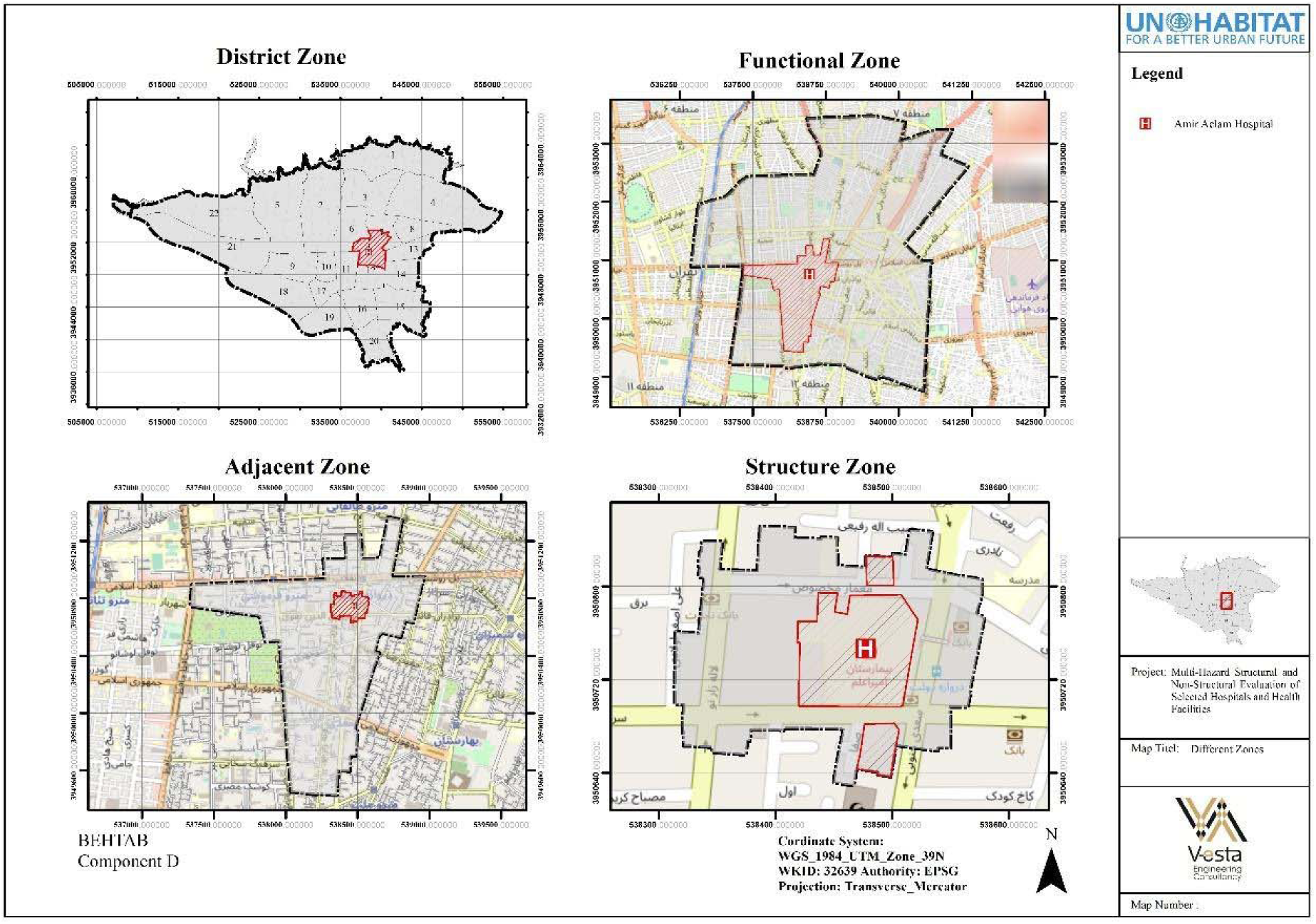
Different Zones Border

### Assessment Framework

We have used a multidimensional framework that simultaneously measures physical and non-physical resilience. Accordingly, a customized assessment framework has been established based on City Resilience Profiling Tool (CRPT) set by the United Nations Office for Disaster Risk Reduction (UNISDR) and available data. We used 36 indicators in 5 components (Physical Attribute, Functional Attribute, Spatial Attribute, Organizational Attributes, and Dynamic Attributes) (**Figure 3**) with a specific scoring system. Ten experts determined the significant coefficients of the components and indices in the proposed scoring system through an AHP technique. Then, the sum of the acquired weights converted to 100 to be more tangible. This system is based on several standards that exist in the global experience, for example, Transit-oriented development (TOD) Standard V3.1 [**32**].

**Figure 3.**
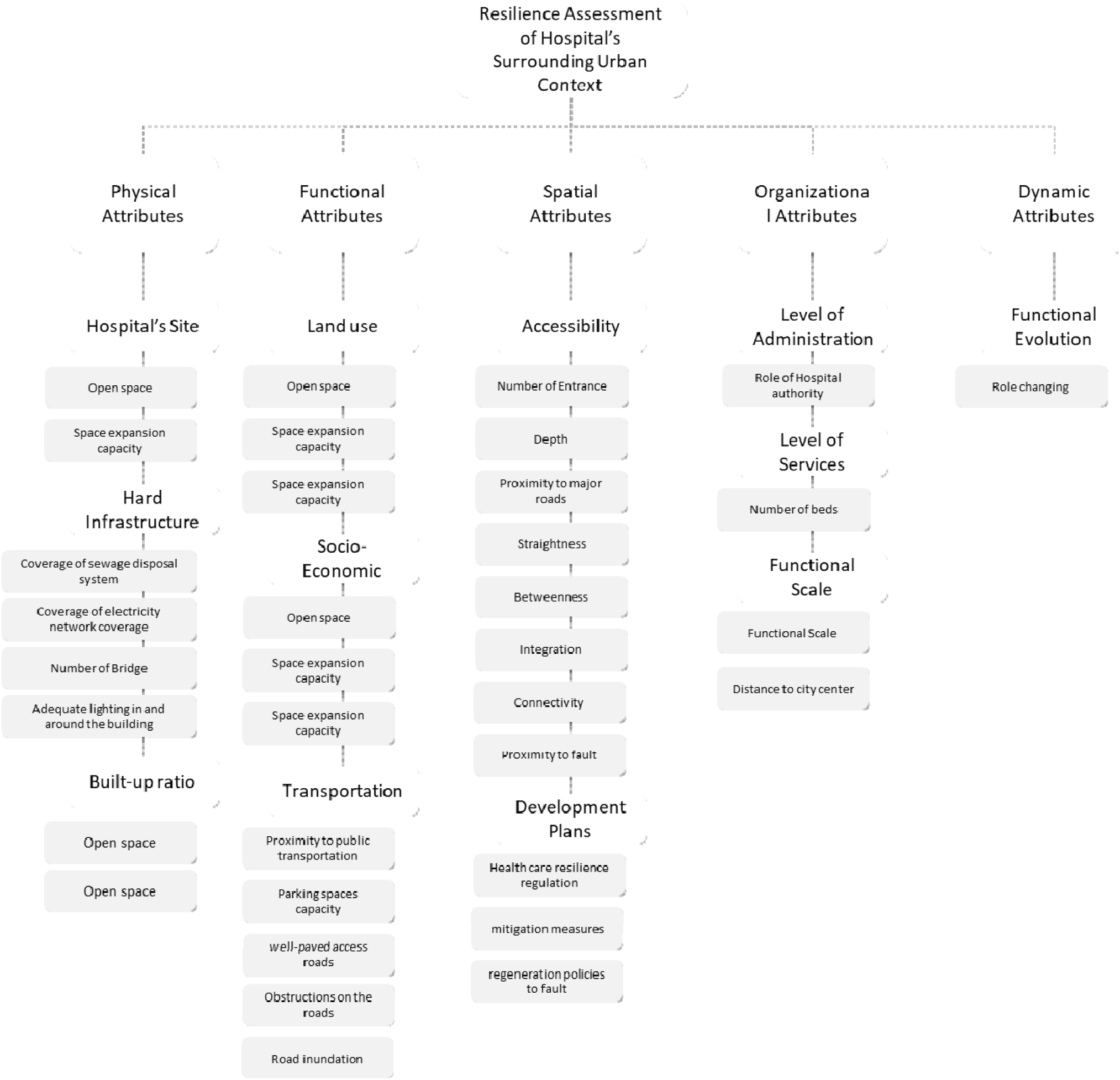
The framework of assessment

After determining the weighting system, the scores of all indicators were calculated based on a specific set of indices (see **Annex 1**). Finally, standardized scores were multiplied by the weights, and the result was obtained. The results justify using the hospital resilience assessments’ results conducted by Zhong et al. [**33**]. In their study, 89.5% identified as having good potential for assessing hospital resilience. Additionally, 20 indicators out of 36 were identified as critical indicators based on experts’ judgment for interpretation of the results. Regarding the significance of the hospitals, it is jointly determined with the authority of the Amir-Alam hospital at the beginning of the assessment that at least 60% of scores for the critical indicators are necessary in order to consider the hospital as a resilient system. The analyses were conducted based on spatial analysis tools in ArcMap 10.2 and space syntax techniques.

### Hospital’s Surrounding Urban Resilience Analysis

#### Component I: Physical Attributes

The first critical component is “Physical Attributes,” consisting of three criteria (Site Plan, Hard Infrastructure, and Built-up Area). The purpose of this criteria is to analyze the physical features of the hospital site and its surroundings in terms of disaster resilience. These criteria comprise various indicators and variables evaluated in the structural zone, adjacent zone, functional zone, or district zone, depending on their nature, ranging from zero to one on a spectrum. The first criterion, “Site Plan,” has two indicators for evaluation, namely “open space” and “space expansion.” Based on general requirements of the standard for planning and design of safe hospitals [**34**], this hospital has not been built accordingly. There is no adequate open space accessible for the gathering because the hospital’s non-built space is 10,000 square meters (**Figure 4**).

**Figure 4.**
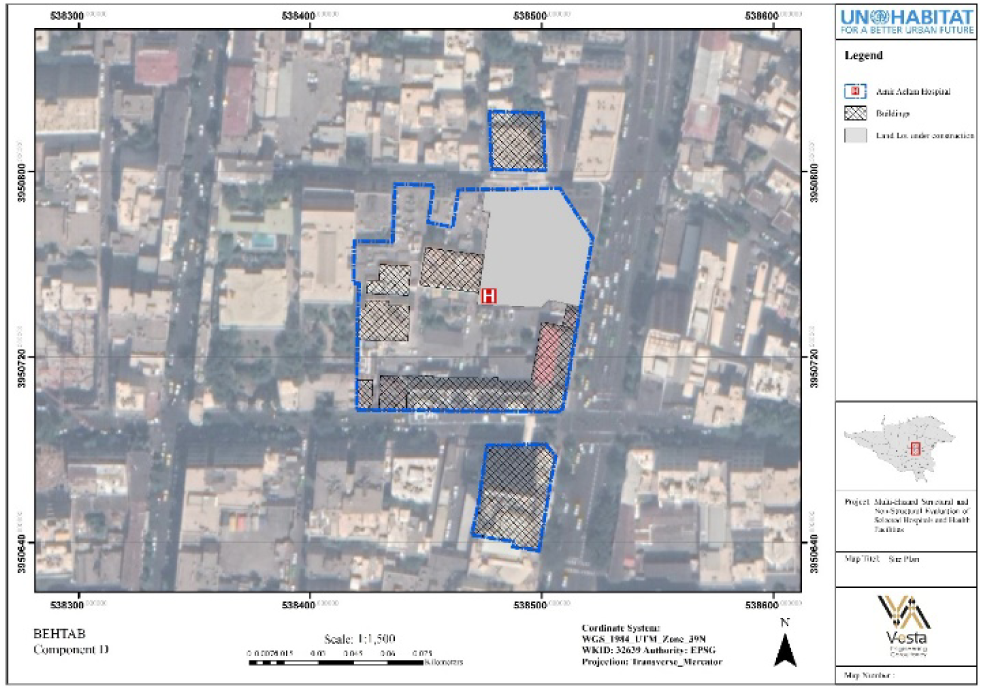
Site plan of Amir-Alam Hospital

The second indicator, “space expansion,” evaluates available space to expand the emergency unit during a crisis. There is insufficient room in this case study to accommodate additional requirements and grow in adjacent areas in the future.

The second criterion, “Hard Infrastructure,” includes lifelines such as “sewage disposal system coverage” and “electricity network coverage” as well as “adequate lighting in and around the building” and “number of bridges” (**Figure 5b**). We employed geographical distribution of underground sewage disposal system and electrical network in the hospital proximity, and there is full coverage of both in the functional zone of the hospital. Furthermore, there are more than two bridges in the hospital’s functional zone, and their demolishing might impair its operation. As a result of the field visit, the lighting around the Amir-Alam hospital is acceptable (**Figure 5a**).

**Figure 5a.**
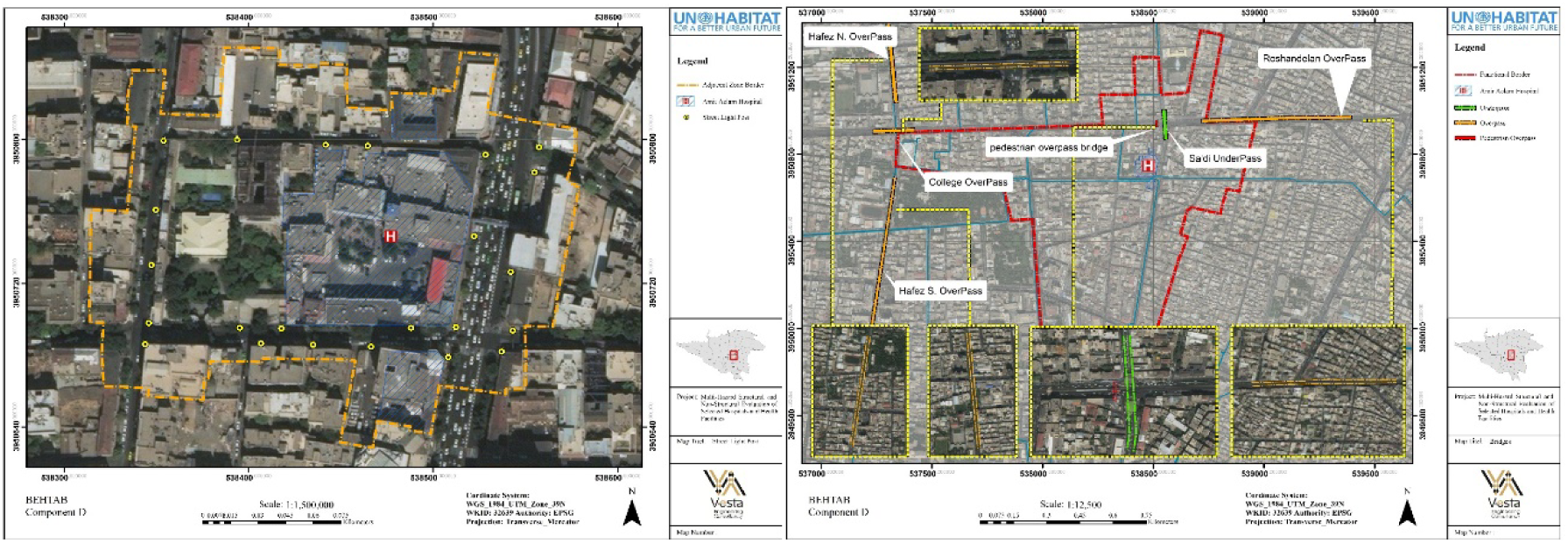
Street Light Post in the adjacent zone of hospital **5b**. Bridges in the functional zone

The third criterion, “Built-up Environment,” includes two indicators: “built-up area ratio” and “floor area ratios (FAR)” (based on Tehran’s master plan). The built-up area compensates for 53% of the total functional area, much more than the recommended ratio for resilience (**Figure 6a**). Furthermore, the proposed FAR on the south and north sides of the hospital functional zone is high, making the region more susceptible through the buildings’ old structure **(Figure 6b**).

**Figure 6a.**
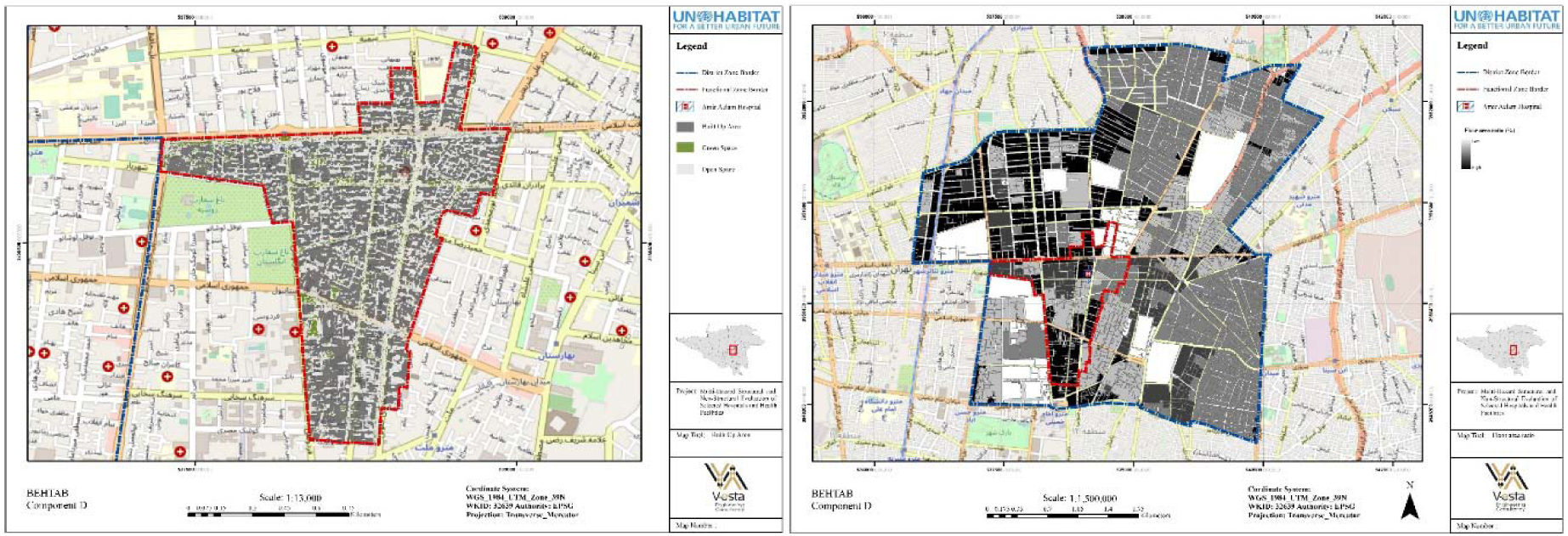
Built-Up Area **6b**. Floor area ratio

#### Component II: Functional Attributes

The second critical component is “Functional Attributes,” including three primary criteria (land-use zoning, socioeconomic, and transportation). Each has some indicators and variables with a range of zero to one as the calculated score.

The first criterion, “land-use zoning,” includes three indicators: “gravity,” “supporting land use,” and “dangerous building.” The Gravity Index, first developed by Hansen [**35**], is still one of the most used spatial accessibility metrics in transportation research. The Gravity Index has been applied to the district zone, with weights based on the building’s population and a query radius of 600 meters (**Figure 7a**) which indicated a high rate of gravity. The current study also investigated the indicator “supporting land use” at two scales of area study: functional and adjacent zone, to see if disaster-supporting land uses were located near the hospital. There are buildings inside the functional zone that serve various functions, such as clinics, pharmacies, mosques, labs, and sports complexes. The adjacent zone, on the other hand, contains just a school. Finally, the indicator “dangerous building” has been considered in the adjoining zone, not including such a building (**Figure 7b**).

**Figure 7a.**
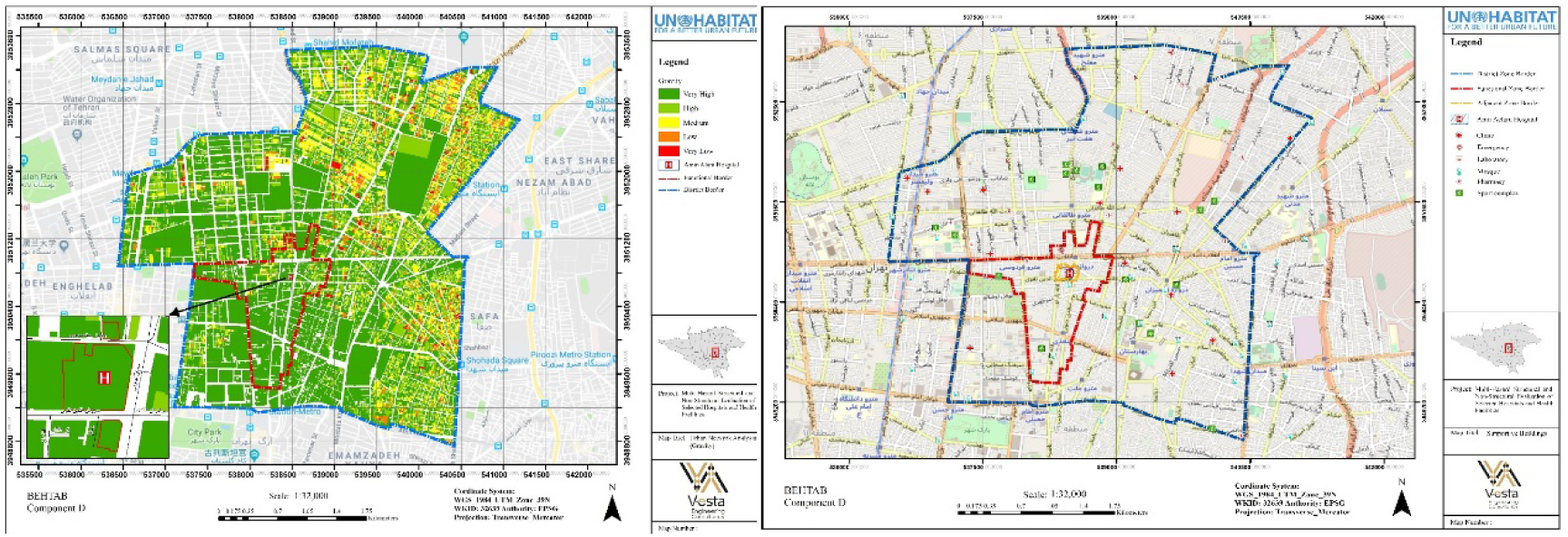
Applying Gravity Index to the dataset in district border **7b**. Disaster supporting building

The second criterion, “Socio-Economic,” includes the indicators “Population density” and “Vulnerable groups” as well as “Literacy ratio” (**Figure 8**). Because more people create additional evacuation challenges, population distribution and density serve as vulnerability indicators [**36**]. As a result, increased population density may lead to weaker resilience. The population density in the hospital functional zone (85 p/ha) is lower than the average population density in Tehran (about 146 p/ha) [**37**]. According to the study, the number of vulnerable individuals (including the elderly (over the age of 65), toddlers (under the age of 6), and disabled groups) in the hospital’s functional zone is low in the majority of blocks. Moreover, because lower education limits the ability to comprehend warning information and access to recovery information [28], it has been used in this case. In comparison to the average of Tehran, 82 blocks out of a total of 112 blocks in the hospital’s functional zone (most of which are situated in the south half of the zone) have a low literacy ratio (**Figure 9**).

**Figure 8.**
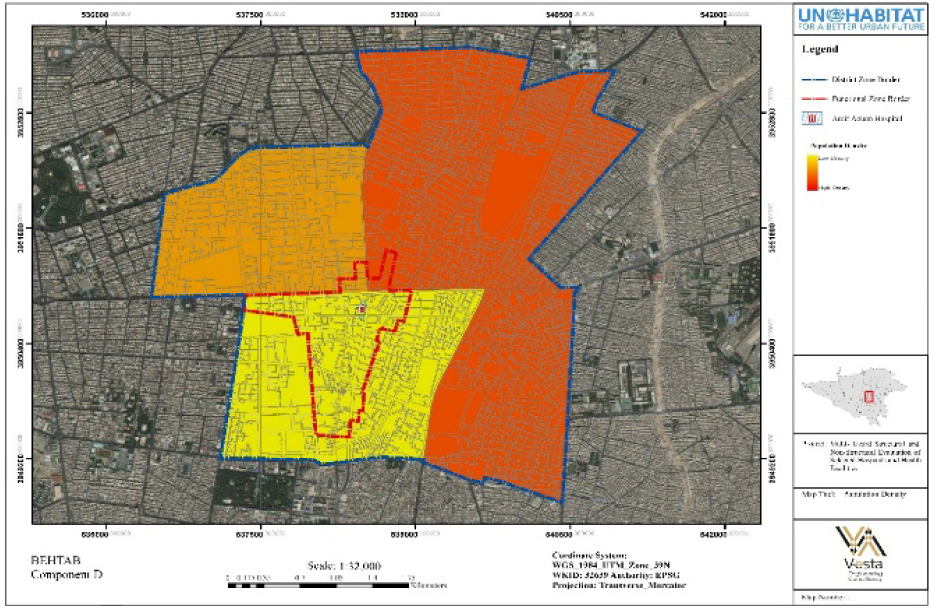
population density

**Figure 9a.**
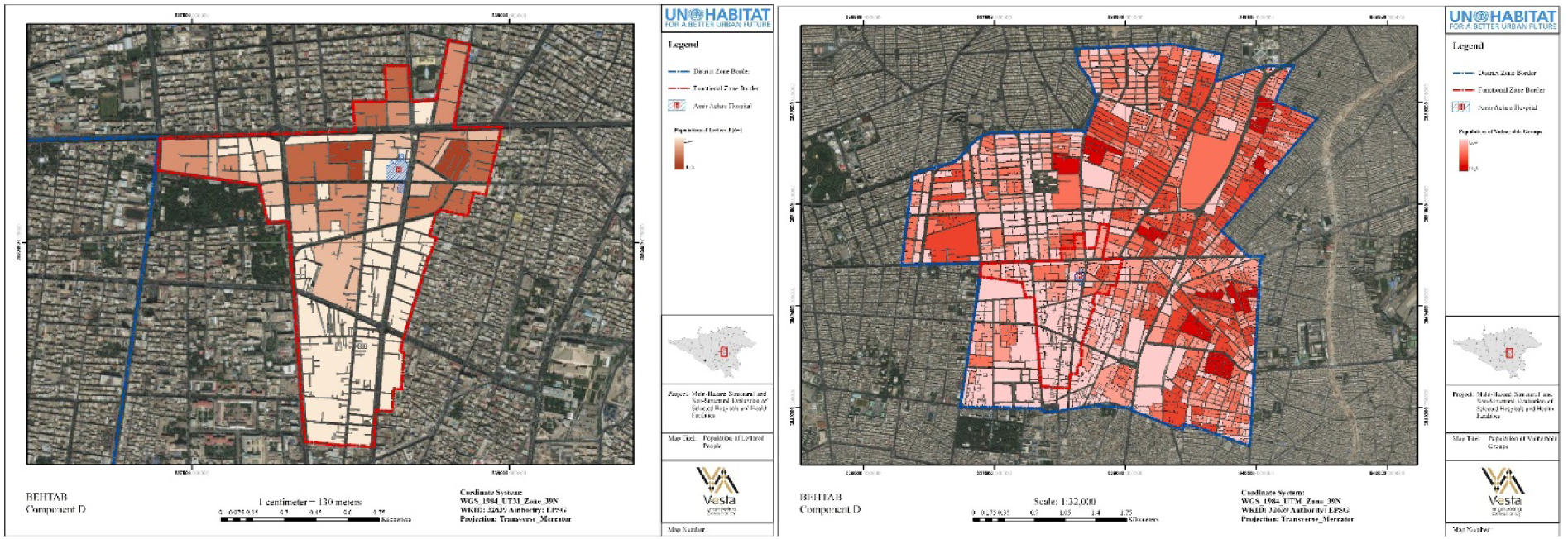
literacy ratio in functional zone of the hospital **9b**. population of vulnerable groups per each

The third criterion, “Transportation,” includes the indicators “proximity to public transportation,” “traffic flow ratio,” “well-paved access roads,” “obstruction,” “road inundation,” and “parking.” The Amir-Alam hospital is close to a metro station (Darwaze-Dolat), a bus station, and a BRT stop. According to Tehran’s most recent annual traffic report [38], the traffic flow ratio in the district zone is medium. Based on field observations, the condition of the paved access road is deemed average in the adjacent zone, since both acceptable and deficient quality may be noticed (**Figure 10a**).

**Figure 10a.**
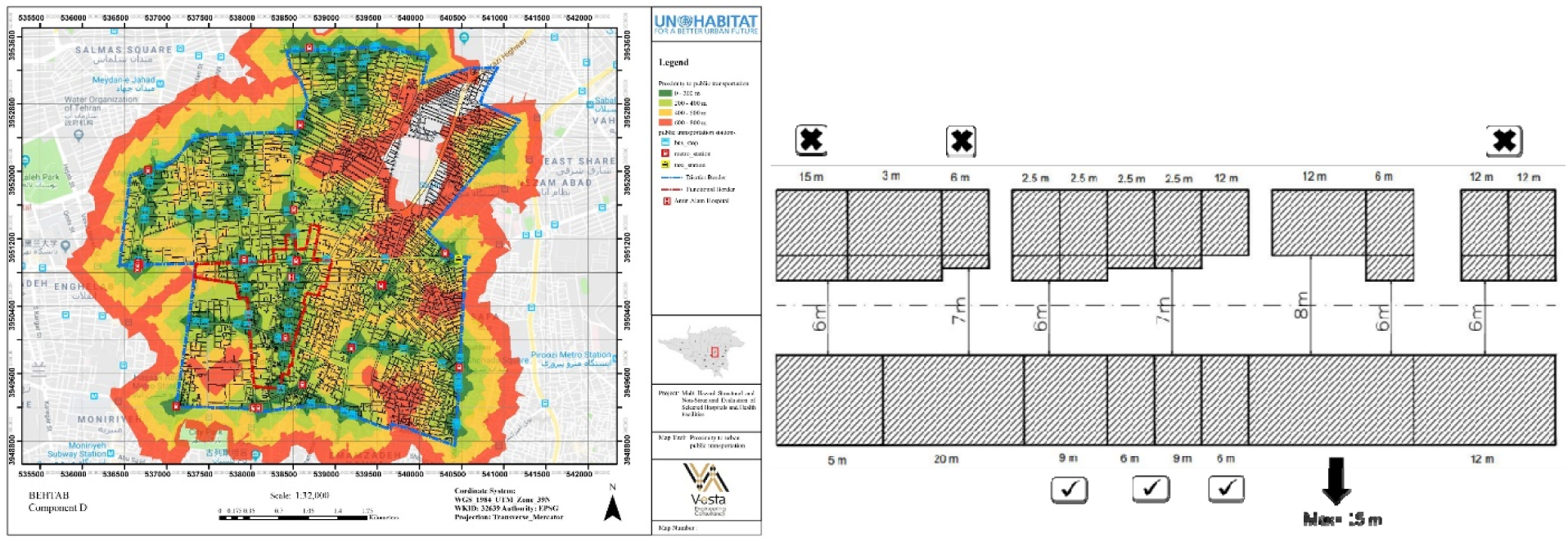
Proximity to public transportation **10b**. the ratio of the street widths to building heights in Memar-Makhsous street

According to Tehran’s master plan [**37**], the maximum height of buildings must be less than three times the width of the road in order to reduce the risk of falling debris on an arterial highway during a crisis. On the “Memar-Makhsous” street, which is the main access road of the hospital, the proportion of street width is calculated. Figure 10b describes the portions of this street that are not suitable in terms of width. Also, according to the flood risk zoning map of Tehran’s master plan, the study area is not in a flood zone; however, surfing water disposal is not operating well in the adjacent zone. Therefore, the road inundation score is rated medium. Furthermore, according to Tehran’s master plan [**37**], the field study reveals that there are enough parking spaces for patients and employees of the hospital.

#### Component III: Spatial Attribute

The third critical component is “spatial attribute,” which includes two criteria, namely: “accessibility” and “development plans.” The main objective of this component is to evaluate the spatial resilience of the hospital within the urban context, particularly accessibility, because it will play a significant role in a post-disaster hospital relief performance.

The first criterion, “accessibility,” is assessed using various indicators to arrive at a precise result. The indicators included in this regard are “number of entrances” of the hospital site, “depth” of surrounding road networks, “proximity of the hospital to major roads,” “straightness” of the roads network, “betweenness” of the hospital within the roads network, “integration, “connectivity” of the hospital in the roads network, and “proximity to fault.” The majority of them are computed using ArcMap 10’s spatial network analysis tool and space syntax technique. According to the general requirement of standard for planning and design safe hospitals [**34**], each hospital building with four entrances would be considered an optimal standard building. This hospital has two entries that can be evaluated with an average score.

Depth is defined as the fewest number of syntactic steps (in topological terms) required to go from one space to another. The hospital has a medium degree of mean depth (**Figure 11a**). The hospital is located less than 200 meters from the major road; Amir-Alam hospital is in good condition for the indicator “proximity to major roads.” The straightness metric shows how near the shortest network distances between two buildings are within a certain radius [**39, 40**]. According to the analysis, the hospital has a medium level of straightness (**Figure 11b**). This indicator should be high for a hospital since it makes it easy for everyone to access. The betweenness of a building calculates how many times it sits on the shortest pathways between pairs of other accessible facilities within the network radius [**41**].

**Figure 11a.**
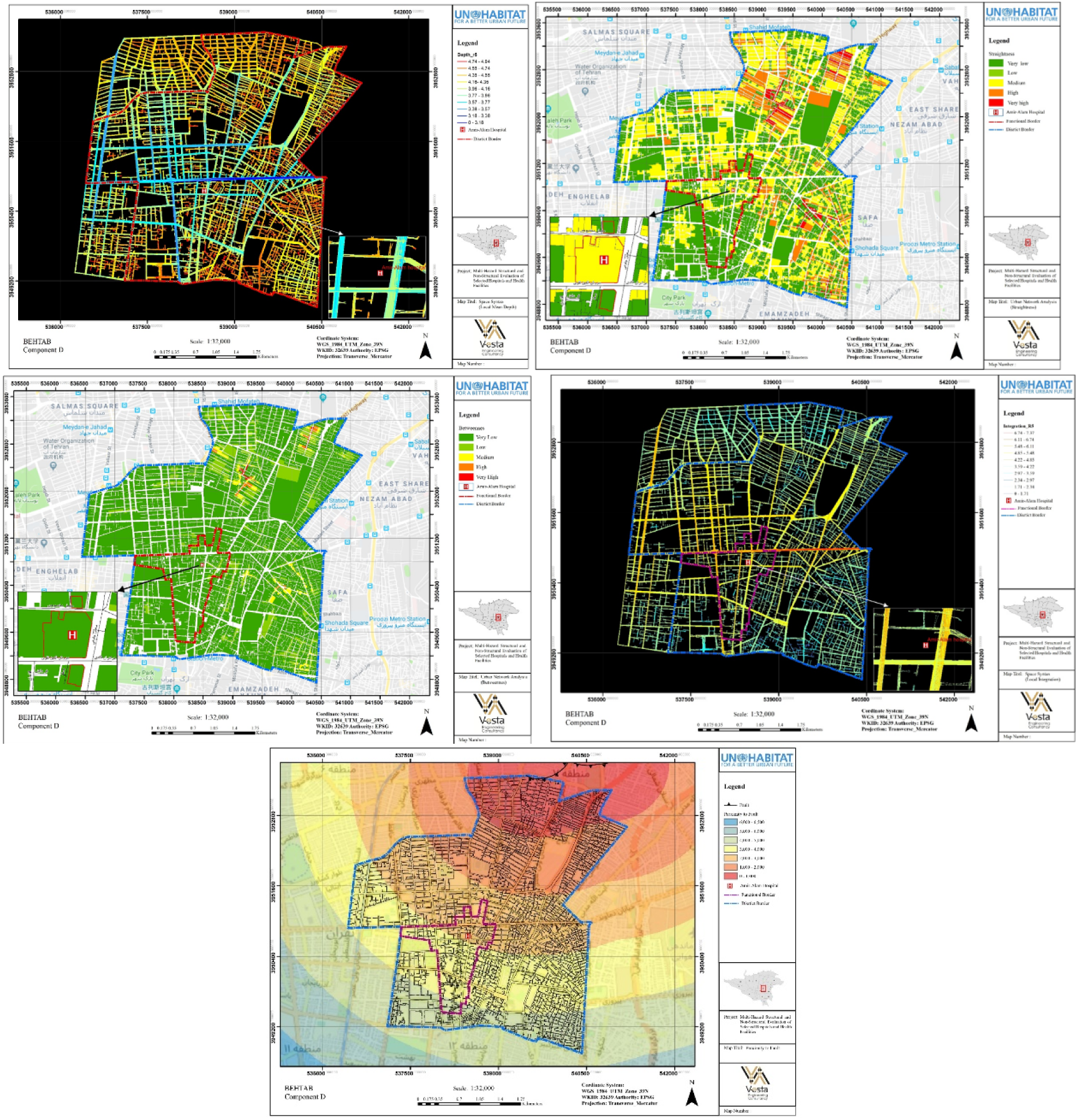
Mean depth of spaces **11b**. Straightness centrality in a 600-meter network radius, weighted by building population **11c**. Betweenness centrality in a 600-meter network radius, weighted by building population in the district area **11d**. Local integration of spaces **11e**. Distance from the fault line

According to the findings, the hospital’s level of betweenness is low, even though the metric should be high for a hospital (**Figure 11c**). Integration, often known as availability, is a variable that describes how a place is connected to other areas in its surroundings. According to the results, the hospital has a high level of local integration (**Figure 11d**), making it easy for everyone to get in. Furthermore, the hospital is approximately 2500 meters away from the fault line (**Figure 11e**), classified as medium risk on the Tehran seismic micro-zonation map.

The second criterion, “development plans,” is included in the study since land-use planning is a powerful tool for decreasing natural disaster losses and building more resilient communities. “Health care resilience regulation,” “mitigation measures,” and “regeneration policies” are three indicators used to assess the amount of adoption of these issues in local planning documents and policies. Based on content analysis techniques, the master plan of Tehran [**37**] was investigated, and the findings revealed that there are no significant regulations on health care resilience, as well as mitigation measures to reduce the vulnerabilities of health care facilities. There are only general guidelines for the regeneration of Tehran’s historic district includes the Amir-Alam hospital.

#### Component IV: Organizational Attributes

In a resilient context, “organizational attributes” refers to any human association for a specific goal, whether official, informal, corporate, or political, and can characterize any organizational scale. We utilized three criteria to examine organizational attributes (including Level of Administration, level of services, and hospital functional scale). The group of administration evaluated analyzing the role of hospital authorities in the community’s decision-making process.

A review of the municipal and local governance profiles reveals a lack of evidence that hospital administration plays an influential role in local government decision-making. The hospital’s serviceability was examined using “number of beds” as an indicator of the second criterion of this component. According to the general requirement of the standard for planning and design of safe hospitals **[34]**, this case study falls within the third level of categories (96 to 300 beds). According to requirements, the service coverage area for these hospitals is about 34337 (square kilometer), which covers 18500 people on average (max= 236381 and min= 141945), and this case study covers about 8000 people, which is by standards. Furthermore, because it is located in district 12 and adjacent to Enqelab Avenue, the Amir-Alam hospital is in Tehran’s core zone.

#### Component V: Dynamic Attributes

Medical and administrative procedures in healthcare are complicated and changing. We assessed the hospital’s dynamic capabilities while considering how the hospital’s function has changed over time. In over two decades, the hospital’s function has shifted from local health care to a district-level hospital in terms of the various services and facilities it delivers. As a result, this hospital receives the maximum score for this component.

### Amir-Alam Hospital’s Urban Resilience Index

The result has revealed that the hospital’s urban resilience score was calculated to be 51.75 out of 100, indicating medium resilience, while, regarding the critical indicators, the score was 20.25 out of 60, which is not acceptable. The table depicts the areas that should address to improving hospital resilience.

## Discussion

In this study, we determined the indicators for assessing hospital resilience using a case-based approach. The proper and timely operation of hospitals is vital in times of outbreaks. Hence, the multi-dimensional assessment of hospital resilience contributes to understanding the drawbacks and challenges in the domain of the disaster risk and eliminating them to mitigate losses and damages [**15**]. The proper performance of hospitals during and post disasters depends on multiple criteria, such as hospital location, building stability, accessibility, and socio-economic indicators. Several existing studies merely addressed the building’s structure and functional services during the crisis [**18**].

Nevertheless, studies that address the influence of inter-settlement relationships in hospital resilience have been rarely found. Thereby, the case-based approach needed to highlight the importance of multiple scalar and dimensional hospital resilience assessment. Like existing studies, we included some indicators that were associated with hospital capacities [**18**]. However, several indicators that describe the multi-scalar operation of the hospital were included in this study.

The literature showed that spatial analysis (e.g., urban access) of hospitals in urban contexts has been less considered. Inter-scalar understanding (interconnected multiple local, metropolitan, regional, national, global scales) of hospital resilience makes a significant difference in risk perception among decision-makers and practitioners. The existing research indicated that safety and preparedness to hazards are merely defined based on building stability and hospital capacity during a crisis. However, this contradicts the resilience concept that aims to protect the continuity of urban systems as one system and mitigate the losses. Resilience does not mean risk reduction by controlling some indicators [**27**]. That is why a comprehensive study of urban systems in risk-prone areas requires the multi-scalar and multi-dimensional understanding and thinking of resilience assessment.

In this study, we designed a framework for spatial risk assessment of the hospital in the urban context. A framework includes multiple indicators, including physical, functional, spatial, institutional, and dynamic. Each of these indicators encompasses sub-indicators that assess the preparedness of the hospital system, such as hospital location and accessibility.

All indicators’ scores are measured by the multiplication of the sub-indicators scores and their allocated weights within the resilience analysis framework. The results reveal that the score of critical indicators was 20.25 out of 60, which means 33.75% of the total score is not acceptable due to the threshold (60%) determined at the beginning of the assessment. So, while the total score of resilience is 51.75, it couldn’t be considered a resilient hospital in the surrounding urban context. After conducting the assessment, all the results were checked and confirmed by the hospital authority in the field as the reliability test.

Furthermore, according to **Table 1**, while the dynamic attribute is in an appropriate condition because of changing hospital role in the urban context, transforming the hospital from a local one to a district-level hospital, the physical attribute is in poor condition. The lack of open space to reclaim it during crisis time and a high built-up area and density rate make the context more vulnerable to disasters. Additionally, there is no sufficient green and open space in the functional boundary. There is just one green space in the 2-minute driving distance from the hospital. The distance of this green space from the hospital is around 1530 meters. The next nearest green space is located within a 3-minute driving distance from the hospital. The network distance of this green space from the hospital is almost 2057 meters. The third and the most significant green space, which is a short distance from the hospital, is 2317 meters from the hospital. In this study, we designed a framework for spatial risk assessment of the hospital in the urban context. A framework includes multiple indicators, including physical, functional, spatial, institutional, and dynamic. Each of these indicators encompasses sub-indicators that assess the preparedness of the hospital system, such as hospital location and accessibility.

**Table 1.**
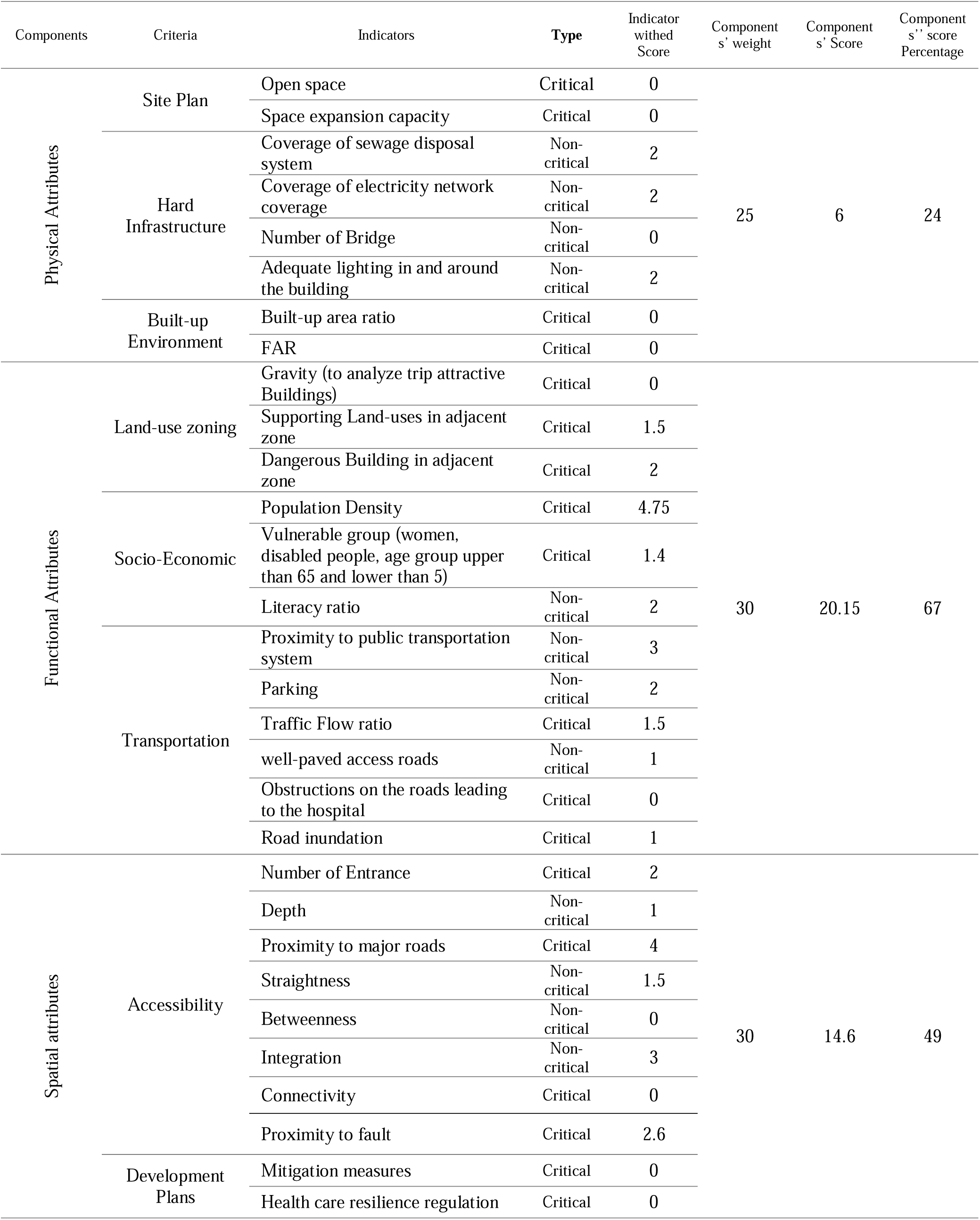
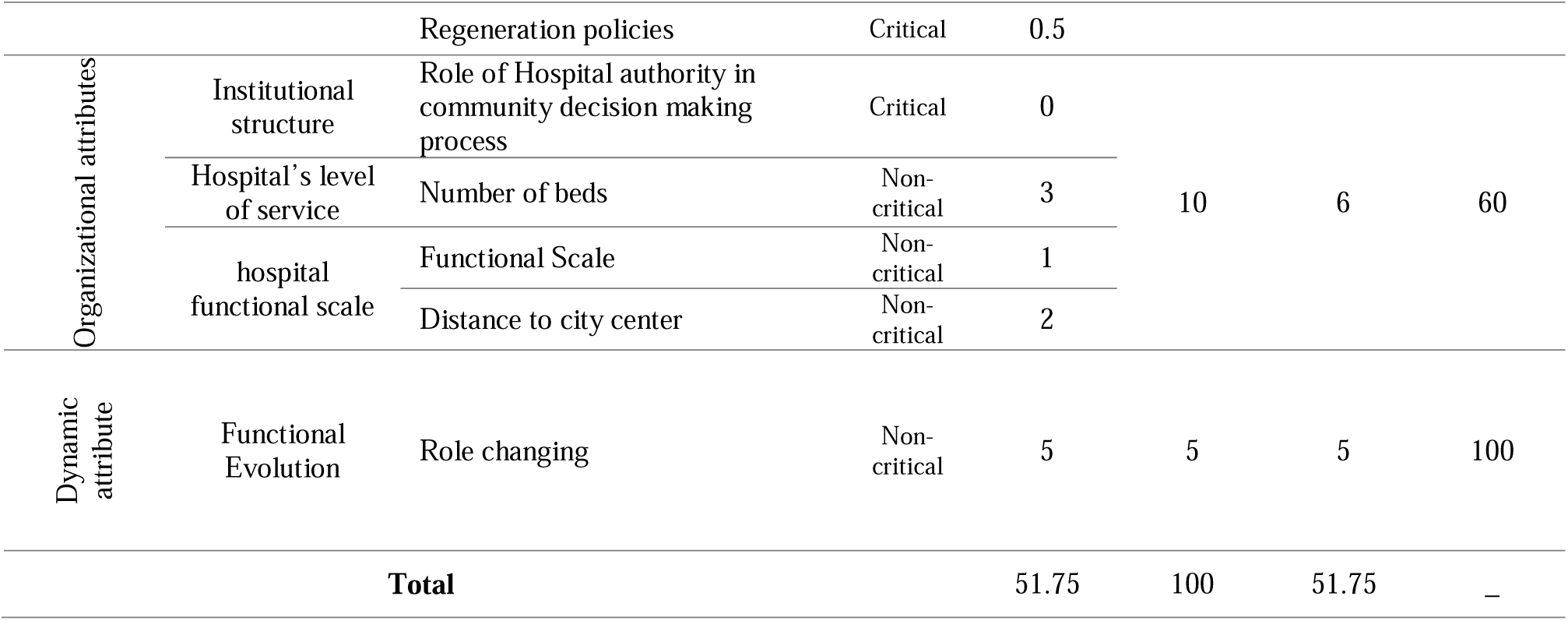
Urban Resiliency index

Besides, functional, spatial, and organizational indicators are in average condition. For instance, in the valuable indicator, the land use of the surrounding area supports the hospital’s operations. In contrast, the urban trips in that area are relatively high and result in traffic congestion, which is inappropriate for the hospital. In the spatial attribute’s component, the indices of accessibility are acceptable, while the development plans do not sufficiently support the hospital’s requirement. In the organization attributes component, while the hospital’s level of services is fair enough, its authority does not actively defend its benefits in the community decision-making process.

Structural indicators are crucial for the building to withstand adverse natural events. Non-structural hands are essential for the daily operations of hospitals and health facilities. If these are damaged, they will not function and may even cause physical injury to patients and personnel. Moreover, functional indicators are essential for the continuous operation of hospitals and health facilities. Identifying and assessing these indicators are necessary at the hospital’s building level and different urban scales, including adjacent functionalities and district zones. Assessing indicators contribute to control risks in hospitals and health facilities while ensuring that they will be resilient, safe, and continue to operate in times of emergency and disaster.

This study provided a list of indicators that must be considered in assessing the vulnerabilities of hospitals and health facilities in the urban context. The next step is to plan for possible actions to enhance the resilience in improving hospital conditions in different boundaries, including structural, adjacent, functional, and district borders. These may include mitigating vulnerability through the following ways:

1. In structural vulnerabilities, some possible measures include:
  ○ Allocating appropriate space (permanent or temporary) in the adjacent and functional boundaries to expand the hospital’s space, especially in times of crisis;
  ○ Renovation of the adjacent buildings of the hospital to reduce the possibility of their collapse during disaster;
  ○ Improving building codes and designs to be more resilient;
  ○ Relocation of the movable elements within and near the hospital’s site to prevent potential obstruction;
  ○ Improving the drainage system, sewage network, and solid waste management.
2. In non-structural vulnerabilities, the focus is to:
  ○ Considering the role and needs of the hospital in the urban development plans as well as community level decision making process;
  ○ Improving the accessibility level of the hospital through traffic management tools;
  ○ Capacity building of community in terms of raising awareness and preparedness against disasters.
3. In reducing functional vulnerabilities, some possible measures include:
  ○ Promoting the pedestrian condition in the functional border in general and in adjacent boundary in particular;
  ○ Considering disabled groups’ access requirements;
  ○ Allocating more supporting land use in the functional zone of the hospital instead of trip attractive or dangerous buildings.

## Limitations

There are three main limitations to this study

1. Our emphasis has been upon the resilience of the urban context that surrounds the hospital, and we did not study the resilience of the hospital’s building and operations. Although we give general information about hospital capacity, we did not collect data associated with operational qualifications of the hospital, such as patient satisfaction, patient waiting time, and hospital performance during the crisis.
2. We overlooked pre-disaster benefit-cost analysis of economic resilience that can contribute to preventing property damage.
3. Hospitals can emit hazardous air pollutants (HAPs) such as mercury and can be responsible for responding to many of the most dangerous effects of pollution and climate change. The environmental factors did not include in the study.

## Conclusion

This study proposed a framework to assess the resilience of a healthcare facility from a new point of view that is generally ignored in previous research. In this study, indicators and sub-indicators assessed the inter-scale interconnection of urban components that surround the hospital. This study quantitatively measured the hospital’s resilience in the urban context that other studies can continue and develop. Moreover, this study suggests using geospatial techniques in healthcare delivery system studies, especially resilience assessment studies.

Further studies should be conducted by developing novel scoring systems in measuring resiliency and adding more indicators to challenge the existing studies. In particular, combining the indicators of a hospital’s building and the surrounding urban context can lead to a more comprehensive assessment. Moreover, vulnerability to disasters varies geographically. Indicators should be validated in future studies by accounting for the dynamic of hazards that differ geographically. Furthermore, assessing the network of health care facilities within a city rather than assessing a single element can contribute to the researches in this field of study. The multi-scalar and multi-dimensional approach in resilience management and assessment can contribute to societies and governments to mitigate hospital vulnerabilities and enhance their performance during and after crises.

## Data Availability

All data is publicly available data

## Acknowledgement

This paper is based on a research project which was supported by UN HABITAT in 2019, funded by the Government of Japan, and fulfilled by VestA Abadgeran Bimarz Engineering Consultancy Ltd, which its area of expertise are structural design and city resilience.

## Conflict of Interest

The authors declare that there is no conflict of interest.

## Notes

### Competing Interest Statement

The authors have declared no competing interest.

### Funding Statement

No funding.

### Author Declarations

No need the IRB approval. Data is secondary data and publicly available

## References

1. Eartuake of damage and of retrofitting techniques in reinforced concrete buildings affected by the 1985 earthquake.

[1] Kanno, T., Koike, S., Suzuki, T. et al. (2019). Human-centered modeling framework of multiple interdependency in urban systems for simulation of post-disaster recovery processes. Cogn Tech Work 21, 301–316.

[2] Mattsson, L.G. & Jenelius, E. (2015). “Vulnerability and resilience of transport systems—A discussion of recent research.” Transp. Res. Part. A: Policy Pract., 81(1), 16–34.

[3] Kamissoko, D. (2013). Decision support for infrastructure network vulnerability assessment in natural disaster crisis situations. Doctorate, University of Toulouse, University of Toulouse 1Capitole.

[4] Rus, K., Kilar, V., & Koren. D. (2018). Resilience assessment of complex urban systems to natural disasters: A new literature review. International Journal of Disaster Risk Reduction 31:311–330.

[5] Zhang, J., Zhang, M. & Li, G. Multi-stage composition of urban resilience and the influence of pre-disaster urban functionality on urban resilience. Nat Hazards 107, 447–473 (2021). https://doi.org/10.1007/s11069-021-04590-3.

[6] Shang, Q., Wang, T., & Li, J.(2020). A Quantitative Framework to Evaluate the Seismic Resilience of Hospital Systems, Journal of Earthquake Engineering, DOI: 10.1080/13632469.2020.1802371.

[7] Salamati Nia, S. P., Kulatunga, U., Udeaja, C. E., & Valadi, S. (2018). Implementing GIS to improve hospital efficiency in natural disasters. The International Archives of the Photogrammetry, Remote Sensing and Spatial Information Sciences, 369-373.

[8] Chen, W., Guinet, A., & Ruiz. A. (2015). “Modeling and simulation of a hospital evacuation before a forecasted flood.” Operations Research for Health Care 4, 36–43.

[9] Zhong S, Clark M, Hou XY, Zang YL, Fitzgerald G. (2014). Development of hospital disaster resilience: conceptual framework and potential measurement. Emerg Med J : EMJ; 31(11):930–8.

[10] Achour, N., Pascale, F., Price, A.D.F., Polverino, F., Aciksari, K., Miyajima, M., et al. (2016). Learning lessons from the 2011 Van earthquake to enhance healthcare surge capacity in Turkey. Environ Hazards-Human Policy Dimen. 15(1):74–94.

[11] Yusoff, N., Shafii, H., & Omar, R. (2017). The impact of floods in hospital and mitigation measures: A literature review. Materials Science and Engineering Conference Series. Johor Bahru.

[12] Ahmadi, A. & Bazargan-Hejazi, S. (2018). 2017 Kermanshah earthquake; lessons learned. J Inju Violence Res.,10(1):1.

[13] UNISDR (2017). Disaster resilience scorecard for cities.

[14] Zhong, S., Clark, M., Hou, X.-Y., Zang, Y.-L., & Fitzgerald, G. (2014a). Development of hospital disaster resilience: conceptual framework and potential measurement. Emerg Med J, 930–938.

[15] Yang, Y., Ng, S.T., Xu, F.J., & Skitmore, M. (2018). Towards sustainable and resilient high density cities through better integration of infrastructure networks, Sustainable Cities and Society, Volume 42, Pages 407–422, ISSN 2210-6707, https://doi.org/10.1016/j.scs.2018.07.013.

[16] Brankov, B., Nenkovic-Rizinic, M., Pucar, M., & Petrovic, S. (2018). “Hospital safety in spatial and urban planning and design– seismic zone in the Kolubara region in Serbia”, Seismic and Energy Renovation for Sustainable Cities.

[17] Djalali, A., Della Corte, F., Foletti, M., Ragazzoni, L., Ripoll Gallardo, A., Lupescu, O., Arculeo, C., von Arnim, G., Friedl, T., Ashkenazi, M., Fischer, P., Hreckovski, B., Khorram-Manesh, A., Komadina, R., Lechner, K., Patru, C., Burkle, F. M., Jr, & Ingrassia, P. L. (2014). Art of disaster preparedness in European union: a survey on the health systems. PLoS currents, 6, ecurrents.dis.56cf1c5c1b0deae1595a48e294685d2f.

[18] Pishnamazzadeh, M., Sepehri, M.M., & Bostadi, B.(2020). An Assessment Model for Hospital Resilience according to the Simultaneous Consideration of Key Performance Indicators: A System Dynamics Approach, Perioperative Care and Operating Room Management, Volume 20, 100118, ISSN 2405 6030, https://doi.org/10.1016/j.pcorm.2020.100118.

[19] Taslimi, M.S., Azimi, A., & Nazari, M. (2021). “Resilience to economic sanctions; case study: hospital equipment cluster of Tehran (HECT)”, International Journal of Disaster Resilience in the Built Environment, Vol. 12 No. 1, pp. 13–28. https://doi.org/10.1108/IJDRBE-06-2018-0024

[20] Khademi Jolgehnejad, A., Ahmadi Kahnali, R., & Heyrani, A. (2020). Factors Influencing Hospital Resilience. Disaster Medicine and Public Health Preparedness, 1–8. doi:10.1017/dmp.2020.112

[21] Narjabadifam, P., Hoseinpour, R., Noori, M. et al. Practical seismic resilience evaluation and crisis management planning through GIS-based vulnerability assessment of buildings. Earthq. Eng. Eng. Vib. 20, 25–37 (2021). https://doi.org/10.1007/s11803-021-2003-1

[22] Cristiano, S., Ulgiati, S., & Gonella, (2021). F. Systemic sustainability and resilience assessment of health systems, addressing global societal priorities: Learnings from a top nonprofit hospital in a bioclimatic building in Africa. Renew. Sustain. Energy Rev. 141, 110765.

[23] Mulyasari, F., Inoue, S., Prashar, S., Isayama, K., Basu, M., Srivastava, N., & Shaw, R. (2013). Disaster preparedness: looking through the lens of hospitals in Japan. International Journal of Disaster Risk Science, 4(2), 89–100.

[24] Zhong, S., Clark, M., Hou, X.-Y., Zang, Y., & FitzGerald, G. (2014b). Development of key indicators of hospital resilience: a modified Delphi study. Journal of Health Services Research & Policy, 1-9. doi:10.1177/1355819614561537

[25] Webb, R., Bai, X., Smith, M.S. et al. (2018). Sustainable urban systems: Co-design and framing for transformation. Ambio 47, 57–77 https://doi.org/10.1007/s13280-017-0934-6

[26] Roggema R. (2020). The Convenient City: Smart Urbanism for a Resilient City. In: Biloria N. (eds) Data-driven Multivalence in the Built Environment. S.M.A.R.T. Environments. Springer, Cham.

[27] Meerow, S., Newell, J. P., & Stults, M. (2016). Defining urban resilience: A review. Landscape and urban planning, 147, 38–49.

[28] Cutter, S. L., Boruff, B. J., & Shirley, W. L. (2003). Social vulnerability to environmental hazards. Social science quarterly, 84(2), 242–261.

[29] Jabareen, Y. (2013). Planning the resilient city: Concepts and strategies for coping with climate change and environmental risk. Cities 31: 220–229.

[30] Coaffee, J., Therrien, M.-C., Chelleri, L., Henstra, D., Aldrich, D. P., Mitchell, C. L., Tsenkova, S., & Rigaud, É. (2018). Urban resilience implementation: A policy challenge and research agenda for the 21st century. Journal of Contingencies and Crisis Management, 26, 403–410.

[31] UN Habitat. (2018). City Resilience Profiling Tool (CRPT). Retrieved January 28, 2020, from http://urbanresiliencehub.org/wp-content/uploads/2018/02/CRPT-Guide.pdf.

[32] ITDP (2018). TOD Standard. Version 3.1.

[33] Zhong, S., Clark, M., Hou, X.-Y., Zang, Y. and Fitzgerald, G. (2015), “Development of key indicators of hospital resilience: a modified Delphi study”, Journal of Health Services Research and Policy, Vol. 20 No. 2, pp. 74–82.

[34] Ministry of health (2013). Standard for planning and design of safe hospitals, standards and general requirement.

[35] Hansen, W.G. (1959). How Accessibility Shapes Land Use. Journal of the American Planning Association, Vol. 25, No. 2, p. 73–76.

[36] Cutter, S. L., Mitchell, J. T., & Scott, M. S. (2000). Revealing the vulnerability of people and places: a case study of Georgetown County, South Carolina. Annals of the association of American Geographers, 90(4), 713–737.

[37] Boomsazegan Ltd. (2007). Tehran’s Master Plan. Ministry of Housing and Urban Development. Iran.

[38] Tehran’s Transportation Organization (2020). Annual traffic report.

[39] Vragovic I., Louis, E. Diaz-Guilera, A. (2005). Efficiency of information transfer in regular and complex networks. physics Review E., 71(026122).

[40] Porta S., Crucitti P., Latora V. (2005). The network analysis of urban streets: a primal approach. Environment and Planning B, Vol. 35, No. 5, p. 705–725.

[41] Freeman L.C. (1977). A set of measures of centrality based on betweenness. Sociometry, 40, p. 35-41.

